# Influence of social determinants of health and county vaccination rates on machine learning models to predict COVID-19 case growth in Tennessee

**DOI:** 10.1101/2021.07.28.21260814

**Authors:** Lukasz S. Wylezinski, Coleman R. Harris, Cody N. Heiser, Jamieson D. Gray, Charles F. Spurlock

## Abstract

The SARS-CoV-2 (COVID-19) pandemic has exposed health disparities throughout the United States, particularly among racial and ethnic minorities. As a result, there is a need for data-driven approaches to pinpoint the unique constellation of clinical and social determinants of health (SDOH) risk factors that give rise to poor patient outcomes following infection in US communities.

We combined county-level COVID-19 testing data, COVID-19 vaccination rates, and SDOH information in Tennessee. Between February-May 2021, we trained machine learning models on a semi-monthly basis using these datasets to predict COVID-19 incidence in Tennessee counties. We then analyzed SDOH data features at each time point to rank the impact of each feature on model performance.

Our results indicate that COVID-19 vaccination rates play a crucial role in determining future COVID-19 disease risk. Beginning in mid-March 2021, higher vaccination rates significantly correlated with lower COVID-19 case growth predictions. Further, as the relative importance of COVID-19 vaccination data features grew, demographic SDOH features such as age, race, and ethnicity decreased while the impact of socioeconomic and environmental factors, including access to healthcare and transportation, increased.

Incorporating a data framework to track the evolving patterns of community-level SDOH risk factors could provide policymakers with additional data resources to improve health equity and resilience to future public health emergencies.

## Introduction

The SARS-CoV-2 (COVID-19) pandemic exacerbated health inequities throughout the United States disproportionately affecting at-risk populations.^1^ Identifying social determinants of health (SDOH) risk factors within US communities that contribute to poor outcomes following infection can improve health equity and strengthen community readiness for future public health emergencies.^2, 3^ Following vaccine rollouts in 2021, we predicted Tennessee COVID-19 case growth using machine learning models and investigated the influence of SDOH factors on COVID-19 incidence to quantify and track opportunities to improve health equity.

## Methods

Our approach combined publicly available COVID-19 testing, vaccination, hospitalization, and death metrics with county-specific SDOH and demographic data.^4, 5^ We employed feature engineering and selection to identify novel county-level predictors such as offset case counts, summed and averaged case growth statistics, and days since the k^th^ COVID-19 case to best capture trends in Tennessee county COVID-19 incidence between February and May 2021. We aggregated data from multiple sources, including the Tennessee Department of Health, John’s Hopkins Coronavirus Research Center, and the U.S. Census database to minimize any implicit bias, and removed or ignored missing values depending on the model type. We trained and tested multiple machine learning models using a grid search approach, including neural networks and tree-based methods, with four to six weeks of historical COVID-19 case data to generate COVID-19 case predictions at thirteen timepoints. We selected optimal models using cross-validation and holdout metrics (e.g., Tweedie deviance, mean absolute error, R^2^).^6^ We analyzed the impact of all features using permutation importance to quantify and rank particular SDOH by their relative influence on COVID-19 case growth predictions.^7^ Finally, we generated linear regression models of county COVID-19 case growth using vaccination rates as regressors to calculate correlation coefficients that quantify the association between local vaccination rates and COVID-19 incidence.

## Results

Machine learning models across all timepoints were more than 90% accurate when comparing model predictions to actual cases (Supplementary Figure 1A & C). The top models demonstrated an average R^2^ value of 0.99, mean absolute error of 0.21, and 0.001 mean Tweedie deviance (Supplementary Figure 1B).

Highly predictive SDOH features changed in importance over time. Categorically, demographic SDOH were most important in February 2021, but socioeconomic and environmental SDOH became increasingly more influential towards May. Health outcome SDOH features remained largely consistent during the study period. Individually, the female and under-18 age demographic features ranked highest in February and then declined while African American poverty and health infrastructure features, such as the number of hospital beds and community provider access statistics, increased in importance by mid-April. Lastly, COVID-19 vaccination data features grew in relative importance by May compared to the other SDOH factors (Figure 1).

**Figure 1.**
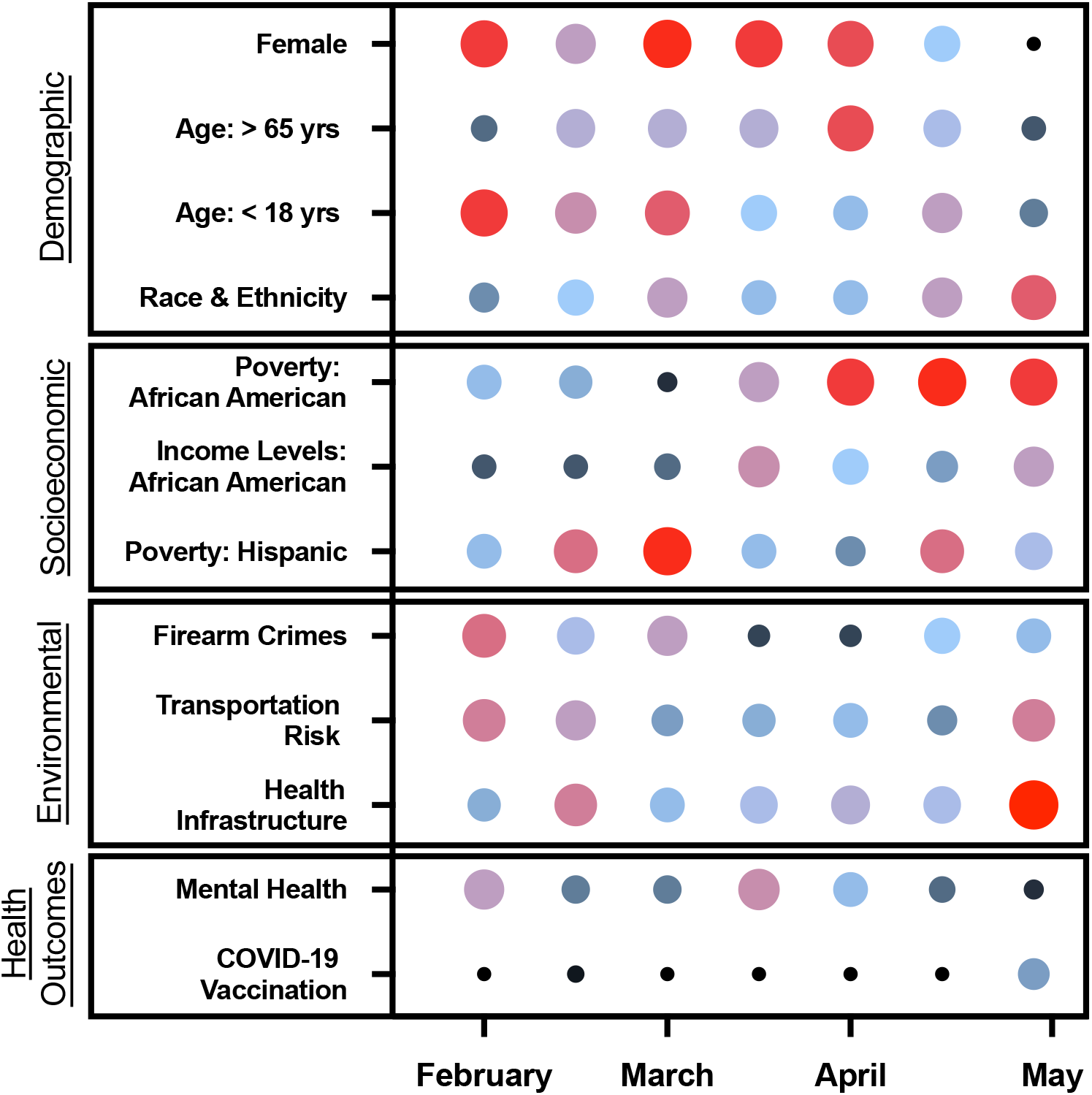
Social determinants of health (SDOH) linked to COVID-19 case growth in Tennessee dynamically shift in importance over time. SDOH include social, physical and environmental factors that impact community health such as age, race, gender, access to transportation, access to primary care and community vaccination rates. Twelve of these SDOH features demonstrated the highest feature importance across all predictive models during the study period. Size and color are used to emphasize SDOH feature importance at each timepoint. Large, red 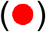 bubbles connote the top ranked SDOH feature while small dark blue 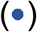 bubbles signify least importance of a given feature at each timepoint. Black bubbles 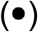 represent the least important feature at each time point compared to the other top ranked SDOH data elements.

As Tennessee vaccination rates increased, counties with the lowest vaccination rates exhibited the highest COVID-19 case growth (Supplemental Figure 2A). Initially, vaccination rates were not correlated with COVID-19 risk, but by mid-March, a statistically significant correlation with low risk of COVID-19 case growth emerged (Supplemental Figure 2B).

## Discussion

Efforts to curtail the health and economic impact of the SARS-CoV-2 pandemic illuminate the need to define specific risk factors that catalyze future case growth, worsen health disparities, and adversely impact the public health response across US communities. Addressing these challenges, we constructed a real-time predictive framework to discover and rank county-level SDOH risk factors that drive machine learning predictions of future COVID-19 case growth (Figure 1).

In Tennessee, we found that communities with rapid vaccine rollout were at lower risk for case growth (Supplemental Figure 2). As vaccination levels began to rise, demographic SDOH features such as age, race and ethnicity declined in relative importance while socioeconomic and environmental risk factors such as poverty, access to transportation and healthcare infrastructure increased significantly. Measures promoting health equity rely on constant assessment of risk mitigation effectiveness. Real-time knowledge of community specific SDOH risk factors empowers healthcare organizations and local governments to improve policy and resource allocation to mitigate outbreaks and enhance resilience to future public health threats.

## Data Availability

The data that support the findings of this study are available from the corresponding author upon reasonable request.

## Acknowledgements

This work was supported by Decode Health, Inc., IQuity Labs, Inc., and grants from the National Institutes of Health (AI124766, AI129147 and AI145505). Dr. Spurlock had full access to all of the data in the study and takes responsibility for the integrity of the data and the accuracy of the data analysis. Dr. Spurlock devised the concept and study design. All authors took part in acquisition, analysis and interpretation of the data along with drafting and revising the manuscript.

