## Supplemental Figure 1 for "Influence of social determinants of health and county vaccination rates on machine learning models to predict COVID-19 case growth in Tennessee"

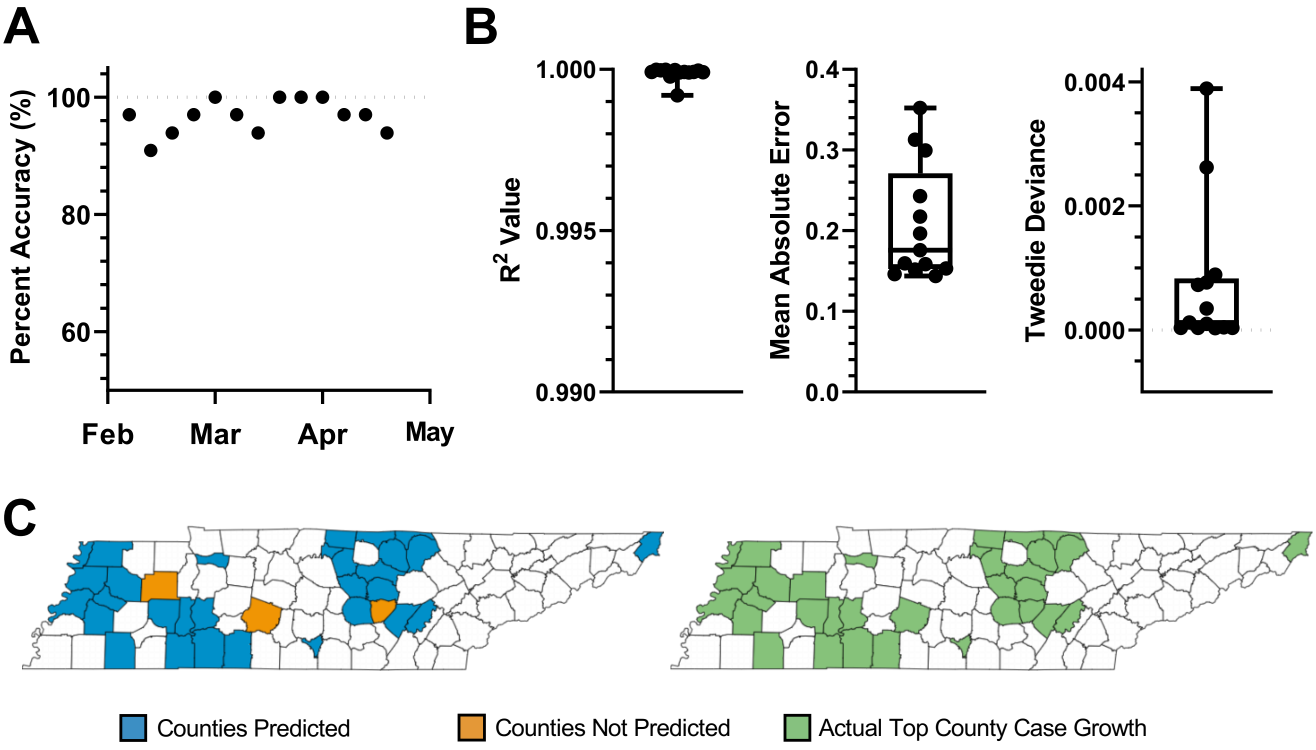


**Supplementary Figure 1. Machine learning models that incorporate historical COVID-19 case growth statistics and SDOH data accurately predict future COVID-19 case growth in Tennessee. (A)** Model accuracy at each time point for predicting counties that will experience the highest COVID-19 case growth normalized to population. We set future population-normalized COVID-19 case growth as the target for predictive modeling and performed a grid search of generalized linear and tree-based machine learning model. Case growth predictions were compared to actual case numbers to determine accuracy. **(B)** Cross-validation metrics of top models at each time point. Shown are $R^2$ values, mean absolute error, and Tweedie deviance. **(C)** Representative illustration of counties predicted (■) or not predicted (■) for future highest case growth versus those that recorded the highest case growth normalized to population at the predicted timepoint. The top third of Tennessee counties with the highest case growth are depicted (■).
