## Supplemental Figure 2 for "Influence of social determinants of health and county vaccination rates on machine learning models to predict COVID-19 case growth in Tennessee"

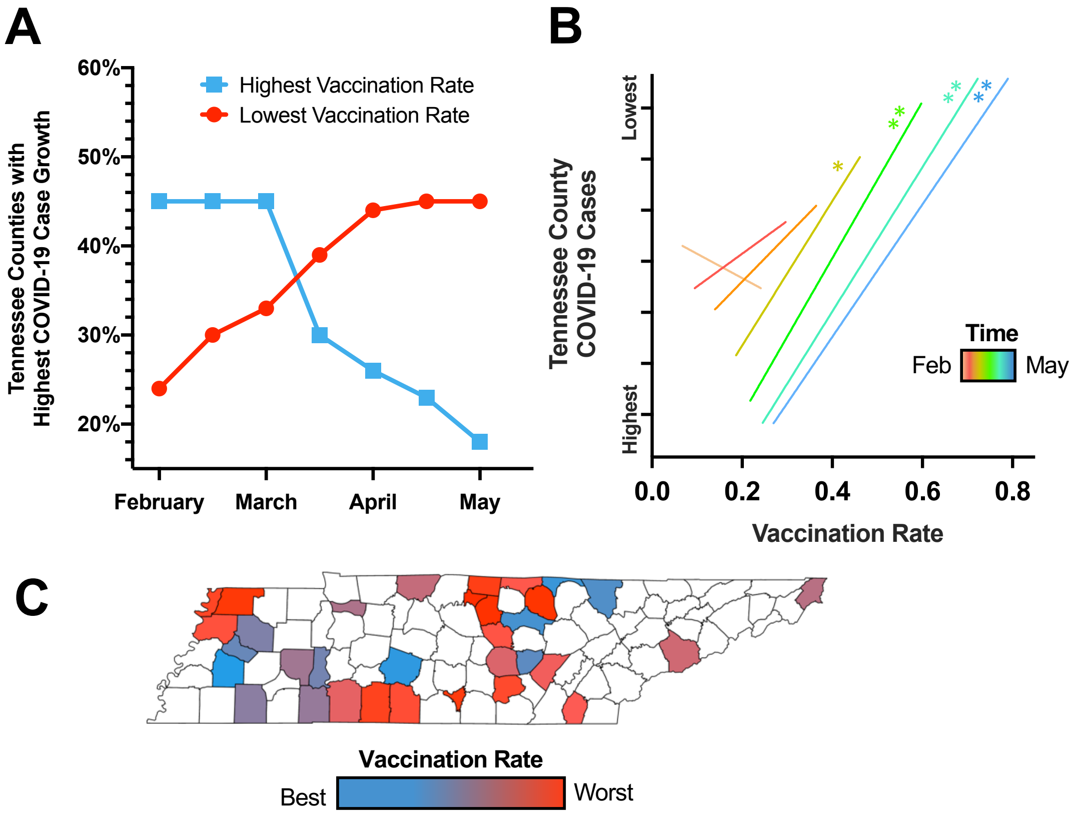


**Supplementary Figure 2. Tennessee county vaccination rates correlate with risk for future COVID-19 case growth. (A)** Counties at each time point with the highest population normalized vaccination rate and lowest vaccination rate are plotted as a proportion of the top third of Tennessee counties with the highest predicted COVID-19 case growth. **(B)** Depiction of the line of best fit for each time point (colored from February to May 2021) using each county's vaccination rate as a regressor for county COVID-19 case count rankings. The slope represents the correlation between county COVID-19 cases and county vaccination rates. The corresponding intercepts depict changes in vaccination rate and COVID-19 cases over time. By the fourth timepoint, vaccination rate is significantly correlated with decreasing COVID-19 case growth. This significance was measured by an ANOVA test performed on the coefficient for vaccination rate. $p$-values less than 0.05 were considered significant: * = $p < 0.05$; ** = $p < 0.01$. **(C)** Heatmap illustration of the top third of Tennessee counties with the highest predicted COVID-19 case growth at the final timepoint overlayed with county vaccination rates. Tennessee counties with the highest COVID-19 case growth risk are enriched for counties with lower overall vaccination rates.
